# D614G Spike Variant Does Not Alter IgG, IgM, or IgA Spike Seroassay Performance

**DOI:** 10.1101/2020.07.08.20147371

**Authors:** Carleen Klumpp-Thomas, Heather Kalish, Jennifer Hicks, Jennifer Mehalko, Matthew Drew, Matthew J Memoli, Matthew D Hall, Dominic Esposito, Kaitlyn Sadtler

## Abstract

Emergence of a new variant of spike protein (D614G) with increased infectivity and transmissibility has prompted many to analyze the potential role of this variant in the SARS-CoV-2 pandemic. When a new variant emerges, there is a concern regarding whether an individual exposed to one variant of a virus will have cross-reactive immune memory to the second variant. Accordingly, we analyzed the serologic reactivity of D614 (original) and G614 variant spike proteins. We found that antibodies from a high-incidence population in New York City reacted both toward the original D614 spike and the G614 spike variant. These data suggest that patients who have been exposed to either SARS-CoV-2 variant have humoral immunity that can respond against both variants. This is an important finding both for SARS-CoV-2 disease biology and for potential antibody-based therapeutics.

## MAIN TEXT

The need to understand the SARS-CoV-2 pandemic has led to the need to understand the rate of infection and acquisition of immunity in the community. The human immune system responds to SARS-CoV-2 infection through a variety of cellular and humoral effectors, including antibodies produced by B cells. Immune antibodies raised against SARS-CoV-2 have been detected that recognize multiple SARS-CoV-2 proteins including nucleocapsid (N), envelope (E), and spike (S) proteins. Antibodies are highly specific for presented epitopes, and mutations in viruses can lead to reductions in immune responses based on acquired immunity to prior viral exposure (Lucas et al., 2001), or even during the course of a single infection (so-called viral escape). It was recently demonstrated by Korber *et al*. through variant tracking that the original SARS-CoV-2 genome spike protein sequence has been supplanted by a changed amino acid at position 614, from D614 to G614 (Korber et al., 2020). Furthermore, evidence suggests that the emergent (and now dominant) G614 virus is more infectious, but a great deal remains to be elucidated including a patient’s potential to be infected by both variants in at once (Grubaugh et al., 2020).

The SARS-CoV-2 spike protein is an outward facing homotrimer presented on the surface of the nucleocapsid that mediates binding to the host cell’s angiotensin-converting enzyme 2 (ACE2). To test for the presence of antibodies against SARS-CoV-2, we and others have developed enzyme-linked immunosorbent assay (ELISA) based seroassays. Our protocol utilizes two types of recombinant primary antigens--full spike ectodomain protein (SARS-CoV-2 S2P) and RBD domain protein (RBD)--as the primary antigens in separate assays for IgG, IgA, and IgM levels (Klumpp-Thomas et al., 2020). The spike 614 position is not within the RBD domain (although, it is represented in full spike ectodomain constructs), and reported seroassays (including ours) are based on the originally observed aspartic acid at position 614 (D614) (Esposito et al., 2020).

The consequence of utilizing D614 spike domain in seroassays is that it could impact the specificity of these assays, given that it is likely most infections in the US are now occurring with the G614 variant of SARS-CoV-2. Knowledge of cross-reactivity is essential to interpreting serosurveys and clinical antibody tests (Hicks et al. 2020). As serosurveys are well underway, we sought urgently to clarify whether serum from recovering/convalescent donors was cross-reactive to both forms of spike protein. Here, we generated a G614 full spike ectodomain construct and incorporated this protein as antigen in an ELISA assay. This spike G614-based assay was compared to the original D614-based assay, and a set of 88 positive samples from a hard-hit (high-incidence) community were applied to both assays.

To evaluate the ability of antibodies developed during SARS-CoV-2 infection to react against both D614 and G614 variants of the spike protein, we measured serologic reactivity via an enzyme linked immunosorbent assay detecting IgG, IgM and IgA binding to full spike ectodomain trimers. Soluble spike trimers were produced that contained a protein sequence identical to the original S-2P spike variant (Wrapp et al., 2020) except for the addition of the D614G mutation. DNA constructs were generated by synthesis (ATUM, Inc.) with gene optimization for expression in human cells and were subcloned into a high-yielding mammalian expression vector driven by a strong CMV51 promoter. Proteins were expressed in Expi293 cells and purified as previously published (Esposito et al). In these vectors, the yields of both D614 and G614 spike proteins were similar (approximately 8-10 mg/L). Both proteins purified in identical fashion, and no difference in protein behavior in analytical size exclusion chromatography was observed, suggesting that both proteins equivalently formed the expected trimeric structures. Using the same ELISA conditions described previously, we tested 88 samples from a high-incidence community and 100 pre-pandemic negative controls (archived pre-2019 sourced from pre-existing NIH study NCT01386424) and compared them to our previously published data for the original D614 spike. We found that sera from donors who tested positive for anti-spike antibodies using D614 spike also tested positive using G614 spike for IgG, IgM and IgA antibodies (Fig. 1a). These data positively correlated for all antibody sub-classes, and all donors who tested positive for D614 antibodies also displayed strong reactivity to G614 spike (Fig. 1b). To further evaluate this correlation, we normalized the IgG values within the linear range of the detector (OD < 3) with the mean of the archival negative controls (Fig. 1c). After running a linear regression and correlation analysis, we found a strong correlation (Pearson R = 0.9735, p < 0.0001) and good fit (R^2^ = 0.9476) between both variants. Furthermore, the slope of the regression line was 1.091 suggesting a 1:1 signal intensity ratio.

**Figure 1:**
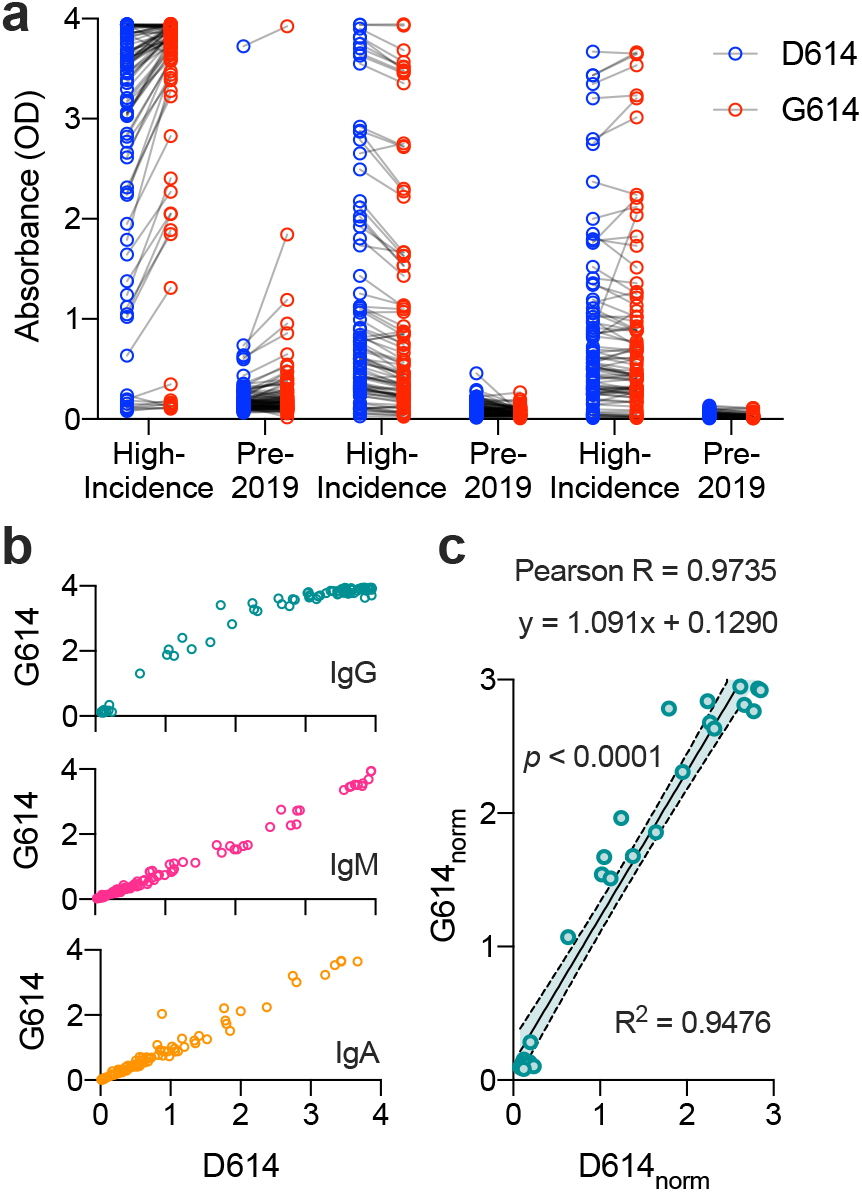
D614G Mutation does not alter antibody binding to either spike variants. (a) Raw absorbance (A450-A650) values for n = 88 donors from a high-incidence community and n = 100 archival pre-2019 donors. (b) Absorbance signals of D614 (original) and G614 (new dominant variant) spike proteins for IgG (teal) IgM (pink) and IgA (orange) antibody classes. (c) Correlation of signal intensity of IgG after normalization of high-incidence samples to mean of archival negative controls (linear range, OD < 3). Linear regression y = 1.091x + 0.1290, R2 = 0.9476, Pearson R = 0.9735, p < 0.0001 (correlation).

A number of seroassays have been published that utilize various spike constructs, and ELISA assays do not provide detection coverage for all possible SARS-CoV-2 antigens, but rather utilize a single protein construct to ascertain seropositivity. Our data show that use of the full spike protein construct should not impact seroassay performance or “miss” seropositive samples. The fact that D614 and G614 both elicited seropositivity is perhaps expected, given that the human immune response is polyclonal (Neurath, 2008; Robbiani et al., 2020). While there may be antibodies produced that recognize spike protein epitopes specific for D614 or G614, these would be among the many antibodies recognizing the SARS-CoV-2 spike protein used in our seroassays (Long et al., 2020). We conclude that human antibody response to SARS-CoV-2 can be detected using D614 or G614 spike protein in ELISA assays.

## Data Availability

Raw data are available from the authors upon request.

## ACKNOWLEDGEMENTS

The authors would like to thank members of the FNLCR Protein Expression Laboratory (William Gillette, Simon Messing, and Vanessa Wall) for support in DNA production and protein purification. This research was supported in part by the Intramural Research Program of the NIH, including the National Institute of Biomedical Imaging and Bioengineering, the National Institute of Allergy and Infectious Disease, and the National Center for Advancing Translational Sciences. This project has been funded in part with Federal funds from the National Cancer Institute, National Institutes of Health, under contract number HHSN261200800001E. Disclaimer: The NIH, its officers, and employees do not recommend or endorse any company, product, or service.

## REFERENCES

1. Esposito, D., Mehalko, J., Drew, M., Snead, K., Wall, V., Taylor, T., Frank, P., Denson, J.P., Hong, M., Gulten, G., et al. (2020). Optimizing high-yield production of SARS-CoV-2 soluble spike trimers for serology assays. Protein Expr Purif 174, 105686.

2. Grubaugh, N.D., Hanage, W.P., and Rasmussen, A.L. (2020). Making sense of mutation: what D614G means for the COVID-19 pandemic remains unclear. Cell Journal Pre-proof.

3. Klumpp-Thomas, C., Kalish, H., Drew, M., Hunsberger, S., Snead, K., Fay, M.P., Mehalko, J., Shunmugavel, A., Wall, V., Frank, P., et al. (2020). Standardization of enzyme-linked immunosorbent assays for serosurveys of the SARS-CoV-2 pandemic using clinical and at-home blood sampling. medRxiv.

4. Hicks, J., Klumpp-Thomas, C., Kalish, H., Shunmugavel, A., Mehalko, J., Denson, J.P., Snead, K., Drew, M., Corbett, K., Graham, B., Hall, M.D., Memoli, M.J., Esposito, D., Sadtler, K. (2020). Serologic cross-reactivity of SARS-CoV-2 with endemic and seasonal Betacoronaviruses. medRxiv.

5. Korber, B., Fischer, W.M., Gnanakaran, S., Yoon, H., Theiler, J., Abfalterer, W., Hengartner, N., Giorgi, E.E., Bhattacharya, T., Foley, B., et al. (2020). Tracking changes in SARS-CoV-2 Spike: evidence that D614G increases infectivity of the COVID-19 virus. Cell In Press.

6. Long, Q.-X., Liu, B.-Z., Deng, H.-J., Wu, G.-C., Deng, K., Chen, Y.-K., Liao, P., Qiu, J.-F., Lin, Y., Cai, X.-F., et al. (2020). Antibody responses to SARS-CoV-2 in patients with COVID-19. Nature Medicine 26, 845–848.

7. Lucas, M., Karrer, U., Lucas, A., and Klenerman, P. (2001). Viral escape mechanisms--escapology taught by viruses. Int J Exp Pathol 82, 269–286.

8. Neurath, A.R. (2008). Immune Response to Viruses: Antibody-Mediated Immunity. Encyclopedia of Virology, 56–70.

9. Robbiani, D.F., Gaebler, C., Muecksch, F., Lorenzi, J.C.C., Wang, Z., Cho, A., Agudelo, M., Barnes, C.O., Gazumyan, A., Finkin, S., et al. (2020). Convergent antibody responses to SARS-CoV-2 in convalescent individuals. Nature.

10. Wrapp, D., Wang, N., Corbett, K.S., Goldsmith, J.A., Hsieh, C.L., Abiona, O., Graham, B.S., and McLellan, J.S. (2020). Cryo-EM structure of the 2019-nCoV spike in the prefusion conformation. Science 367, 1260–1263.

